# Analyzing the Usage of the GIM-COVID-19 Long-Term Sequelae Rapid Access to Consultative Expertise (RACE) Line

**DOI:** 10.1101/2024.01.28.24301875

**Authors:** Saniya Kaushal, Peter Birks, Jesse Greiner, Adeera Levin, Michelle Malbeuf, Zachary Schwartz

**Affiliations:** Providence Health Care Research Institute; Graduation: Spring 2025, Medical Student Research Assistant with Providence Health Care Research Institute; The University of British Columbia Faculty of Medicine; General Internal Medicine Doctor (GIM), Nephrologist, Administrative Lead for Post-COVID-19 care in Fraser Health; The University of British Columbia Faculty of Medicine; General Internal Medicine Specialist, Medical Director of Post-COVID-19 Recovery Clinic at St. Paul’s Hospital; The University of British Columbia Faculty of Medicine; Head of the Division of Nephrology at the University of British Columbia, Nephrologist, Executive Director of the BC Renal Agency; Providence Health Care Research Institute and Provincial Health Services Authority; Program Manager of the Post-Covid-19 Interdisciplinary Clinical Network, PHSA; The University of British Columbia Faculty of Medicine; General Internal Medicine Specialist, Physician Lead for Post-COVID-19 Recovery Clinic at Vancouver General Hospital

**Keywords:** Post-COVID-Interdisciplinary Clinical Care Network (PC-ICCN), Long-Term Sequelae of COVID-19, Rapid Access to Consultative Expertise (RACE) line

## Abstract

Real time access to guidance for physicians has been offered through Rapid Access to Consultative Expertise (RACE) in British Columbia (BC) for the past 12 years. ^1^ In the context of the novel coronavirus (COVID-19), the service for RACE was expanded to include a Long-COVID RACE line. ^1^ We report here the types and frequencies of questions asked to General Internal Medicine (GIM) experts in Long-COVID, by general practitioners in BC. 149 calls over an 11-month period were tracked by GIM experts, and analysis of the call themes was undertaken. These calls mainly involved consults regarding the post-infection COVID-19 symptoms being experienced by Primary Care Practitioners’ patients. Respiratory symptoms were the leading type of symptoms reported, with shortness of breath, cough, fatigue, and fevers being the most common, respectively. This data will be used to inform future resource utilization and provide insights on the usage of the Long-COVID RACE line.

## Introduction

As the novel coronavirus (COVID-19) pandemic continues to unfold, observations of long-term symptoms experienced by COVID-19 patients continue to be recorded. Patients experiencing residual symptoms, such as shortness of breath, cough, and fatigue, for months post-infection, are being diagnosed with Post-Acute Sequelae of SARS-CoV2 (PASC). ^2^ In this study, data regarding PASC patients collected through the General Internal Medicine (GIM) – COVID-19 Long-Term Sequelae division of the provincial Rapid Access to Consultative Expertise (RACE) line, was tabulated to identify trends in the long-term progression of COVID-19 symptoms. Findings of this study will offer information to improve the provincial patient care and long-term support for future COVID-19 patients, while offering insights on the usage of the GIM-COVID-19 Long-Term Sequelae RACE line.

As of June 27, 2022, the novel coronavirus (COVID-19) pandemic has affected over 544 million people worldwide. ^3^ The virus responsible for the novel coronavirus (COVID-19) is the severe acute respiratory syndrome coronavirus 2 (SARS-CoV-2).^2^ Between its detection in December 2019 and June 27, 2022, the COVID-19 virus has infected 3.94 million Canadians.^3^ British Columbia accounts for 373,974 of these cases. ^4^ Although over 369,000 British Columbians have since recovered from their acute COVID-19 illness, many are still experiencing residual symptoms for months or longer. ^4^ These patients have been deemed to be suffering from Post-Acute Sequelae of SARS-CoV2 (PASC). ^5^ This syndrome has many other names too, including Long-COVID and Long haulers; it is heterogeneous in presentation.^2, 5^ This condition’s identification, management, and long-term prognosis remain under investigation.

In British Columbia, Canada, the Post-COVID-Interdisciplinary Clinical Care Network (PC-ICCN) was developed to support the best outcomes for patients recovering from symptoms following COVID-19 infection through research, education, and clinical care. ^6^ One of the clinical resources within the PC-ICCN included the establishment of the GIM – COVID-19 Long-Term Sequelae division of the provincial RACE line. ^6^ This resource provides immediate (<2 hours) specialist advice to general practitioners caring for patients with long-term sequelae of COVID-19 infection. The provincial RACE line has existed in BC since 2010 and provides access to immediate specialist advice/consultation across the province. ^1^

The COVID-GIM-Post-Infection Care RACE line is answered by a dedicated group of General Internal Medicine Specialists with an interest and experience with acute and chronic COVID. ^1^ The guidance provided by the Internal Medicine physicians includes diagnostic investigations, management, and navigation of these complex patients.

This report presents an analysis of the types and frequencies of calls made to the Post-Covid Care division of the RACE line. This analysis enables the identification of trends in patient presentations, primary care practitioner concerns, and related questions. This data informs on the development of education, tools, and care plans which improves the quality of care and long-term support for COVID-19 patients and their health care providers.

## Methods

This is a Quality Improvement (QI) study evaluating the COVID-GIM-Post-Infection Care RACE line data from its launch in August 2020 to June 2021. This RACE data is comprised of the documented exchanges between Primary Care Practitioners (PCPs) and the General Internal Medicine Specialists.

149 RACE line call medical notes were systematically reviewed to extract data regarding the variables of interest: patient demographics (age, sex, region) and types of queries related to COVID-19 (acute symptoms, subacute symptoms, chronic symptoms, vaccination inquiries, miscellaneous questions). The data from these calls were tabulated for analysis. Six calls were excluded from this study because they were too vague to draw conclusive findings. The remaining 143 calls were used to observe trends in age, sex, geographical location, types of queries, timing, and symptoms of COVID-GIM-Post-Infection Care RACE line patients between August 2020 and June 2021.

For RACE calls regarding patient symptoms, we examined the reported symptoms according to the time post-COVID infection. This was predetermined as 0-2 weeks following diagnosis to represent acute COVID-19 symptoms, 2-12 weeks following diagnosis to represent subacute COVID-19 symptoms, and 12+ weeks following COVID-19 diagnosis to represent chronic COVID-19 symptoms. The relative frequencies of types of symptoms reported to the COVID-GIM-Post-Infection Care RACE line were analyzed and compared.

We also analyzed the reasons for calls to the RACE line by time period within the pandemic. Firstly, we described the type of calls received during the different COVID-19 “waves” as occurred in BC, including August to December 2020, January 2021 to March 2021, and April 2021 to June 2021. Secondly, we assessed the reasons for the call before and after the availability of COVID-19 vaccines. We hypothesized that the reason for calls would vary depending on the time period during the pandemic.

## Results

### Demographics

Figure 1 represents the ages of patients about whom PCPs called the RACE line between August 2020 and June 2021. The data indicates the most common age group of RACE line patients as being between the ages 40-49 years old. Of the 113 patients whose ages were recorded, 28.3% of patients fell into this age category (32).

**Figure 1.**
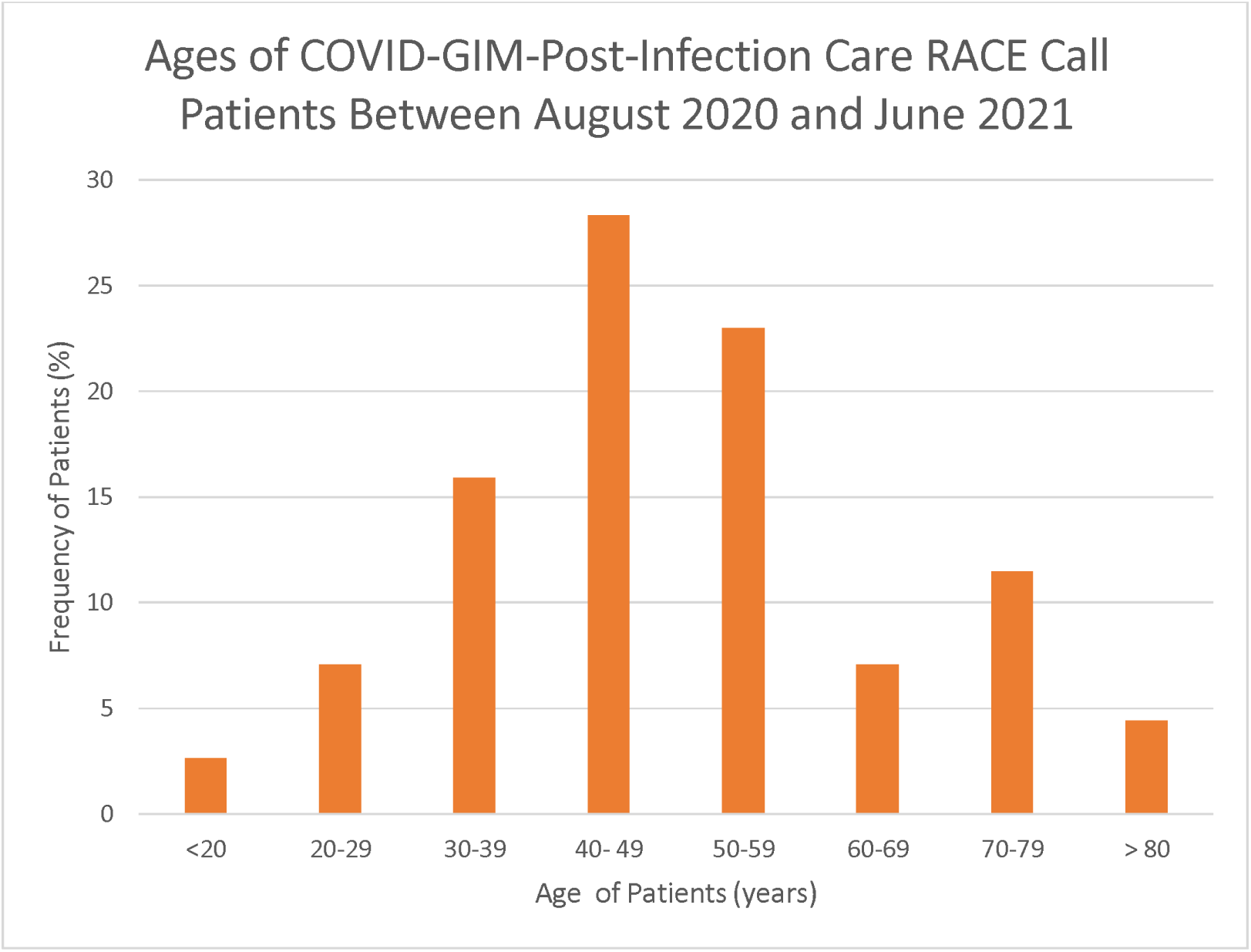
Ages of COVID-GIM-Post-Infection Care RACE Call Patients Between August 2020 and June 2021

The sex of the RACE line call patients are indicated in Figure 2. There was a significantly larger population of female patients than male patients. Of the 143 patients recorded, 91 patients were females, and 52 were males. Therefore, females made up 63.6% of the COVID-GIM-Post-Infection Care RACE line patients, while males made up 36.4% of the patients between August 2020 and June 2021.

**Figure 2.**
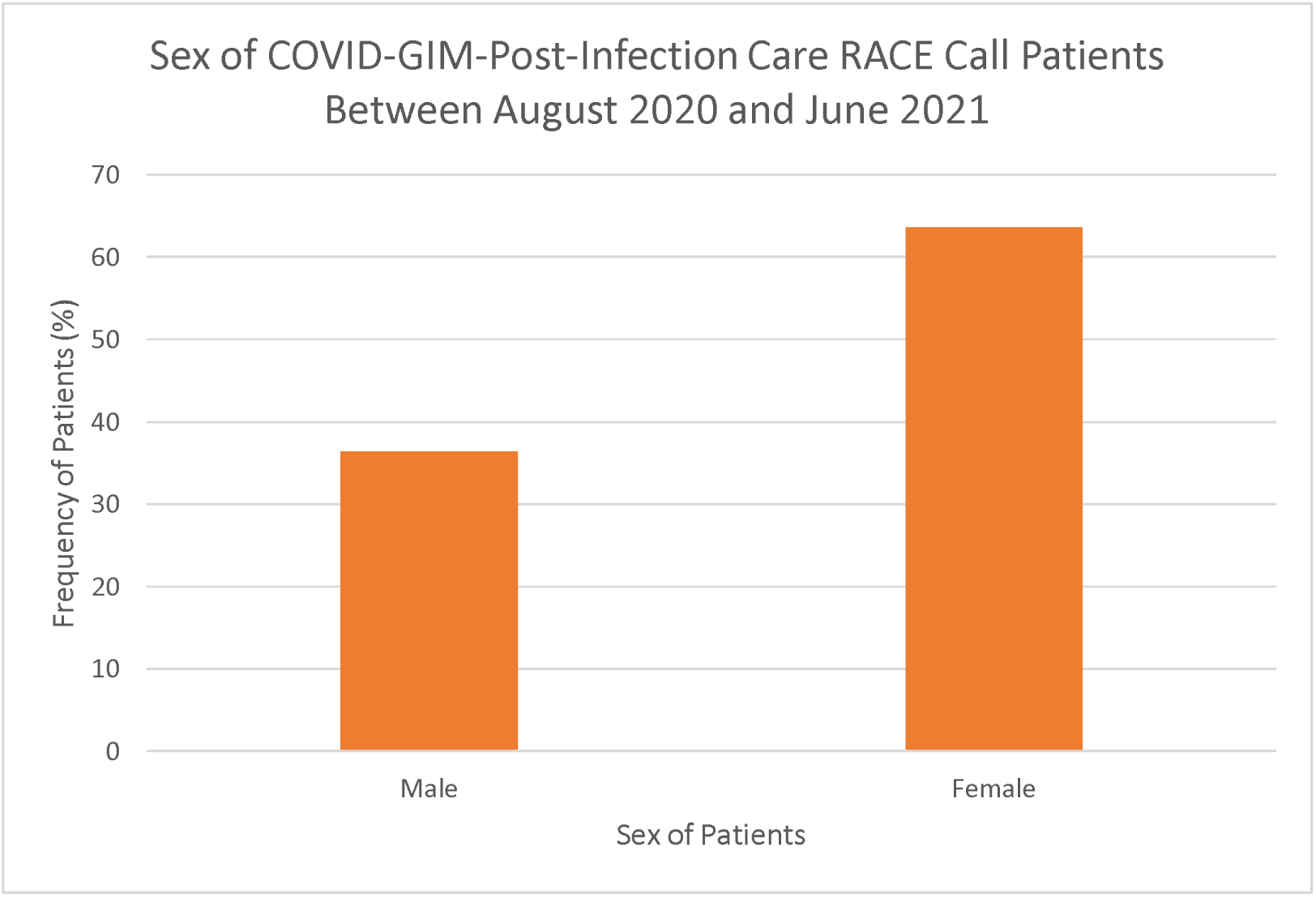
Sex of COVID-GIM-Post-Infection Care RACE Call Patients Between August 2020 and June 2021

Of the 143 calls analyzed, 83 provided information regarding the geographical locations of the Family Physicians and Nurse Practitioners seeking specialty advice from the RACE line’s COVID-GIM-Post-Infection Care division for their patients. This data is represented in Figure 3. As shown, the most frequent geographic locations for the RACE line patient’s healthcare providers were in the Greater Vancouver region (35) and Fraser Valley region (29). These two regions made up 77.1% of the RACE line calls, with Greater Vancouver alone counting for 42.2% and the Fraser Valley counting for 34.9%.

**Figure 3.**
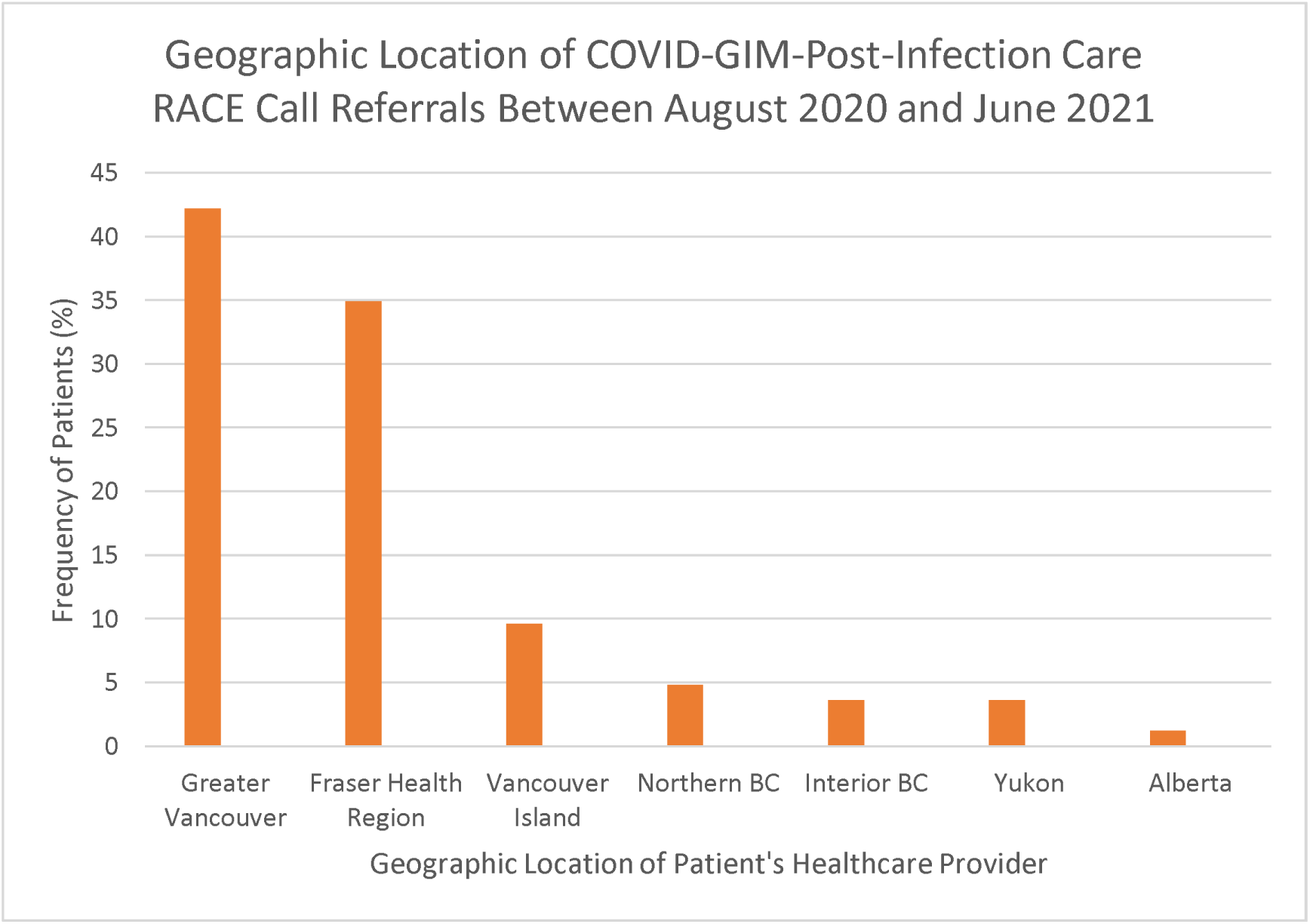
Geographic Location of COVID-GIM-Post-Infection Care RACE Call Referrals Between August 2020 and June 2021

### Types of Queries

Figure 4 shows the types of COVID-19 related queries that were received by the COVID-GIM-Post-Infection Care RACE-Line. The data demonstrates that subacute (2-12 weeks following diagnosis) symptoms (52) and vaccination queries (29) were the most common RACE line call matters, making up 34.9% and 19.5% of all call types respectively.

**Figure 4.**
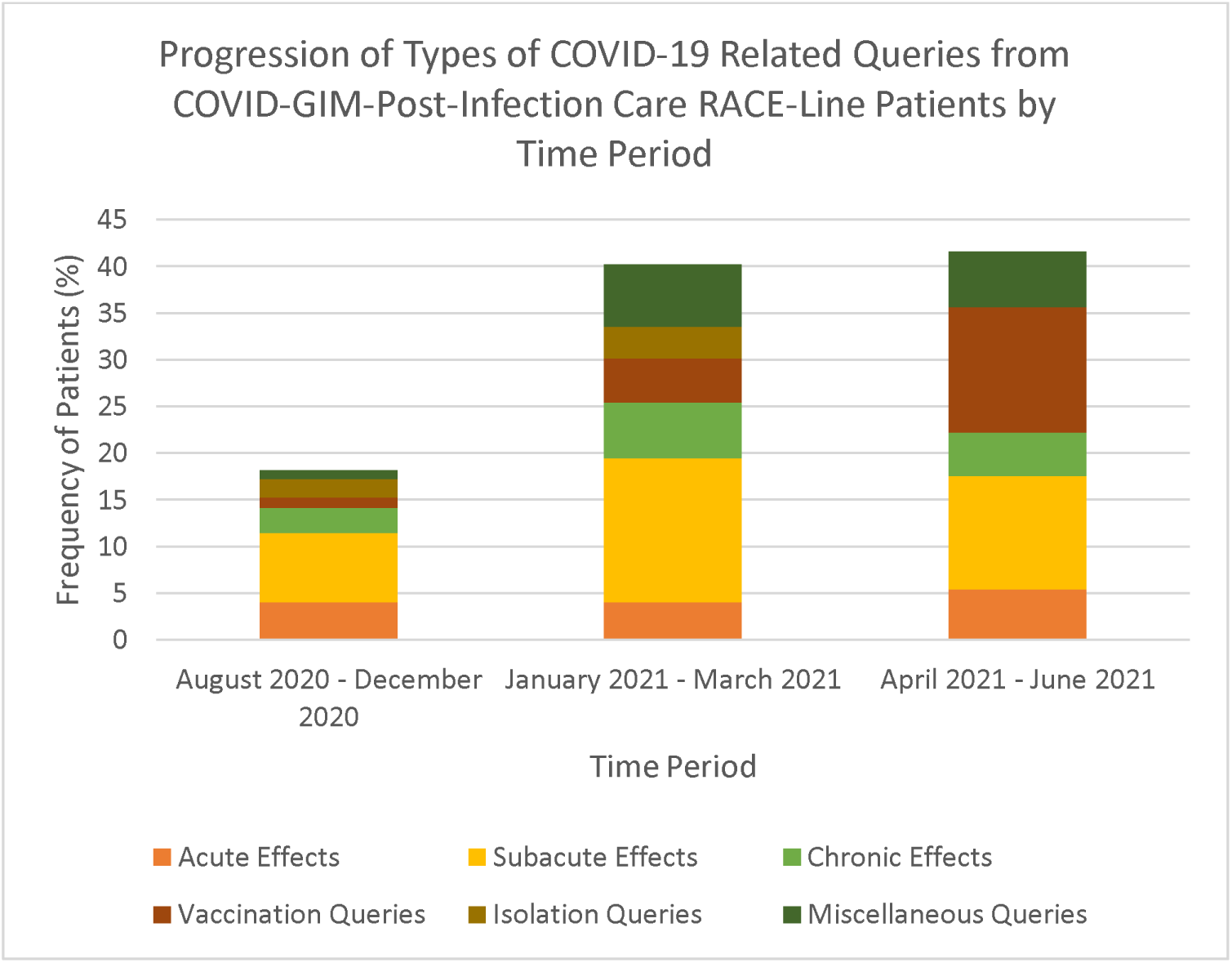
Progression of Types of COVID-19 Related Queries from COVID-GIM-Post-Infection Care RACE-Line Patients by Time Period Examples of miscellaneous queries included questions regarding potential COVID-19 implications due to pre-existing health conditions, and medication/treatment inquiries.

### Timing of Queries

Figure 4 portrays that there was a much larger frequency of RACE line calls to the COVID-GIM-Post-Infection division between the January 2021 - March 2021 (60) and April 2021 – June 2021 (62) time periods, in comparison to the earlier August 2020 – December 2020 period (27). There was a much larger number of queries following the introduction of the COVID-19 vaccine (122) in comparison to during the time period prior (27). Queries made following the COVID-19 vaccine made up 81.9% of overall queries.

### Symptoms

Symptoms reported to the COVID-GIM-Post-Infection Care RACE line, between August 2020 and June 2021, were compiled to provide the data represented by Figure 5. Figure 5 represents the calls which reported symptoms present 2-12 weeks following COVID-19 diagnosis (subacute), or 12+ weeks following COVID-19 diagnosis (chronic).

**Figure 5.**
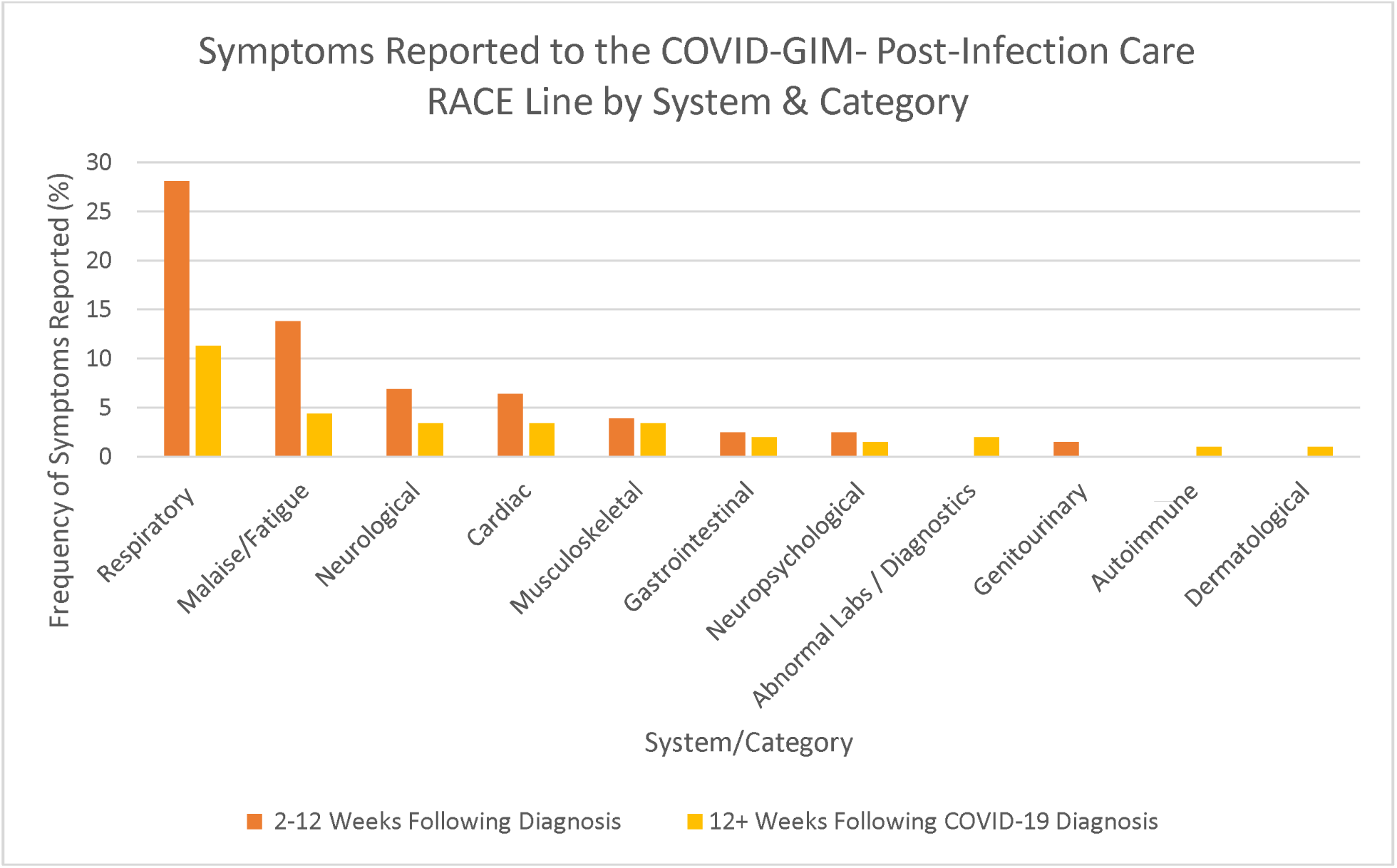
Symptoms Reported to the RACE Line by System/Category

Questions about specific symptoms varied by time post-infection, but the most frequent symptoms across time periods included shortness of breath, cough, and fatigue. Overall, 92 calls focused on symptoms, and within these calls, there were 253 total symptoms reported. Shortness of breath was the most reported symptom, being identified in 35 calls (38.0%). Cough, fatigue, and fevers were the other highest reported symptoms recorded among COVID-GIM-Post-Infection Care RACE line patients, accounting for 28.3%, 25.0%, and 23.9%, respectively. The most common symptoms reported 12 weeks post-infection were shortness of breath, chest pain, and fatigue.

Figure 5 organizes the symptoms reported according to body system. Our findings demonstrated respiratory, malaise/fatigue, and neurological symptoms were the most common categories of post-infection symptoms resulting in calls to the RACE line. Respiratory symptoms made up 39.4% of reported symptoms. Respiratory symptoms reported included cold, coryza, cough, hypoxemia, lung infiltrate, nasal congestion, rhinorrhea, shortness of breath, sore throat, sputum, and wheezing. Shortness of breath, cough, and hypoxemia were the most persistent respiratory symptoms listed, respectively. Neurological symptoms described by PCPs regarding their patients included dizziness, light-headedness, numbness, vertigo, and paresthesia, respectively.

## Discussion

This report provides information on the usage of the COVID-GIM-Post-Infection Care RACE line, specifically between August 2020 to June 2021. Our data suggests that the COVID-GIM-Post-Infection Care RACE line has been a utilized resource for PCPs, especially in the Greater Vancouver and Fraser Health regions. The data obtained indicate that the frequency of calls to the COVID-GIM-Post-Infection Care RACE line has increased throughout the pandemic. Although this RACE line is accessible across the province, there was minimal uptake outside the two aforementioned regions, as seen in Figure 3. This may reflect ‘burden’ and populations affected, or an underutilization by more sparsely populated regions. Further analysis of unmet need vs not needed is required so that we can ascertain if different strategies for awareness of the RACE line outside of populace areas is required. This study highlights the need to understand mechanisms by which GPs learn about new RACE lines, and this resource in particular.

Observable trends in the data indicated the most common age group of RACE line patients as being between the ages 40-49 years old (28%). Females made up 63.6% of the COVID-GIM-Post-Infection Care RACE line patients between August 2020 and June 2021. Interestingly, these demographics are also representative of those mostly likely to be referred and seen in Post-Covid Recovery Clinics (PCRCs) across the province too. Thus, information from clinicians to PCP is based on relevant experience in a similar population.

RACE line subject matters involved acute, subacute, and chronic effects experienced by the patients. Respiratory, malaise/fatigue, and neurological symptoms were the most common categories of post-infection symptoms reported to the RACE line. This data can be used to identify gaps in PCPs knowledge in the diagnosis and management of persistent symptoms following COVID-19, and has informed the development of educational resources for health care practitioners. Adopting these care plans would significantly improve the quality and aid provided by future RACE-line calls in this division. Therefore, increasing the accessibility of resources on managing these identified symptoms should be a focus moving forward. This is an initiative that is currently being implemented by the provincial PCRC websites. Other topics of interest include that RACE line calls also frequently centered around vaccinations, isolation periods, the impact of pre-existing health conditions on COVID-19 manifestation, and anti-viral treatments. This is despite regular bulletins and updates to MDs from provincial health bodies, infectious disease specialists and PHO. This highlights key areas of uncertainty and opportunities for clearer messaging to health care practitioners.

This study examines routinely standardized and collected information generated from RACE calls over an 11-month period of time, where a small number of dedicated individuals were answering calls. To our knowledge, this is the only provincial Post-COVID RACE line set up. The analysis of themes was organized prior, and tools were developed allowing for future assessment of ongoing themes and calls. Another strength is that of the 149 RACE line calls made to the COVID-GIM-Post-Infection Care division during the time period being investigated, only 6 were excluded due to unclarity. Therefore, 96.0% of all applicable calls were used to generate the findings of this QI study. This corresponds to a study which provides a greater overview of the COVID-GIM-Post-Infection Care RACE line calls than one with a lower percentage of population representation. This is indicative of sampling validity, and it translates to a greater elimination of design or inclusion bias. Prejudice in this study was also greatly eliminated as tabulation was performed by an outside party with no pre-existing relationships to the subjects of the studied RACE line calls.

Limitations of this study include that the pre-existing medical conditions of PCPs’ patients were not taken into consideration when tabulating post-infection symptoms reported. Pre-existing conditions may have exacerbated the prevalence of some symptoms of post-acute COVID-19 patients. Additionally, there is no clear differentiation in the severity of symptoms described by PCPs regarding their patients and no standardized approach to measure these symptoms, on RACE line calls. It is also not known if other specialist RACE lines were called for individuals with more specific organ system symptoms (eg. respirology, cardiology, psychiatry). Therefore, this study’s data may reflect a smaller number of true RACE calls for post-COVID.

## Conclusion

The RACE line’s COVID-GIM-Post-Infection Care division was introduced to provide PCPs with prompt medical advice from General Internal Medicine Specialists, regarding chronic COVID. Over the course of this pandemic, this resource has been utilized by health care professionals to improve timely access to care and it has provided support for the appropriate delivery of high-quality care by the PCP. This study investigated the COVID-GIM-Post-Infection Care RACE line calls between August 2020 and June 2021, revealing many trends in data. These calls mainly involved consults regarding the post-infection COVID-19 symptoms being experienced by PCPs’ patients. Respiratory symptoms were the leading type of symptoms reported, with shortness of breath, cough, fatigue, and fevers being the most common, respectively.

Moving forward, RACE calls can be monitored in situations of emerging diseases to better inform and educate community physicians to common complaints that patients are presenting with.

## Data Availability

All data produced in the present work are contained in the manuscript.

## Acknowledgements

The authors of this QI thank all PCPs who utilized the COVID-GIM-Post-Infection Care RACE line, thereby providing the data which was tabulated in this study.

## Notes

### Competing Interest Statement

The authors have declared no competing interest.

### Funding Statement

This study did not receive any funding.

### Author Declarations

Ethics committee of UBC Providence Health Care Research waived ethical approval for this work. Under Article 2.5 of the Tri Council Policy Statement, QA/QI activities are not subject to institutional ethical review.

## References

1. What Is RACE? RACE Rapid Access To Consultative Expertise. http://www.raceconnect.ca/about-race/what-is-race/. Accessed January 6, 2022. [Online Resource]

2. Naming the coronavirus disease (COVID-19) and the virus that causes it. World Health Organization. Retrieved from: https://www.who.int/emergencies/diseases/novel-coronavirus-2019/technical-guidance/naming-the-coronavirus-disease-(covid-2019)-and-the-virus-that-causes-it. Accessed May 12, 2022 [Online Resource]

3. Hannah Ritchie, Edouard Mathieu, Lucas Rodés-Guirao, et al. Coronavirus Pandemic (COVID-19). Our World in Data. Retrieved from: https://ourworldindata.org/coronavirus. Accessed May 12, 2022 [Online Resource]

4. BC COVID-19 Data. BC Centre for Disease Control. Retrieved from: http://www.bccdc.ca/health-info/diseases-conditions/covid-19/data. Accessed May 12, 2022 [Online Resource]

5. Proal AD, VanElzakker MB. Long COVID or Post-acute Sequelae of COVID-19 (PASC): An Overview of Biological Factors That May Contribute to Persistent Symptoms. Front Microbiol. 2021;12:698169. Published 2021 Jun 23. doi:10.3389/fmicb.2021.698169

6. About the PC-ICCN. Provincial Health Services Authority. Available from: http://www.phsa.ca/our-services/programs-services/post-covid-19-care-network/about. Accessed May 12, 2022. [Online Resource]

